# Deep Phenotypic Analysis of Psychiatric Features in Genetically Defined Cohorts: Application to XYY Syndrome

**DOI:** 10.1101/2022.08.29.22279329

**Authors:** Armin Raznahan, Srishti Rau, Luke Schaffer, Siyuan Liu, Ari M. Fish, Catherine Mankiw, Anastasia Xenophontos, Liv S. Clasen, Lisa Joseph, Audrey Thurm, Jonathan D. Blumenthal, Dani S. Bassett, Erin N. Torres

**Affiliations:** Section on Developmental Neurogenomics, Human Genetics Branch, National Institute of Mental Health Intramural Research Program, Bethesda, MD, USA; Center for Autism Spectrum Disorders and Division of Neuropsychology, Children’s National Health System, Washington DC, USA; Neurodevelopmental and Behavioral Phenotyping Service, Office of the Clinical Director, National Institute of Mental Health Intramural Research Program, Bethesda, MD, USA; Departments of Bioengineering, Electrical & Systems Engineering, Physics and Astronomy, Neurology and Psychiatry University of Pennsylvania, Philadelphia, PA, USA; Santa Fe Institute, Santa Fe, NM USA

**Keywords:** Behavioral Phenotyping, Neurogenetics, Symptom Networks, Sex Chromosomes, Deep Phenotyping: Adaptive Function

## Abstract

**Background:** Recurrent gene dosage disorders impart substantial risk for psychopathology. Yet, understanding that risk is hampered by complex presentations that challenge classical diagnostic systems. Here, we present a suite of generalizable analytic approaches for parsing this clinical complexity, which we illustrate through application to XYY syndrome.

**Method:** We gathered high-dimensional measures of psychopathology in 64 XYY individuals and 60 XY controls, plus additional interviewer-based diagnostic data in the XYY group. We provide the first comprehensive diagnostic description of psychiatric morbidity in XYY syndrome and show how diagnostic morbidity relates to functioning, subthreshold symptoms, and ascertainment bias. We then map behavioral vulnerabilities and resilience across 67 behavioral dimensions before borrowing techniques from network science to resolve the mesoscale architecture of these dimensions and links to observable functional outcomes.

**Results:** Carriage of an extra Y-chromosome increases risk for diverse psychiatric diagnoses, with clinically impactful subthreshold symptomatology. Highest rates are seen for neurodevelopmental and affective disorders, and a lower-bound of <25% of carriers are free of any diagnosis. Dimensional analysis of 67 scales details the profile of psychopathology in XYY, which survives control for ascertainment bias, specifies attentional and social domains as the most impacted, and refutes stigmatizing historical associations between XYY and violence. Network modeling compresses all measured symptom scales into 8 modules with dissociable links to cognitive ability, adaptive function, and caregiver strain. Hub modules offer efficient proxies for the full symptom network.

**Conclusion:** This study parses the complex behavioral phenotype of XYY syndrome by applying new and generalizable analytic approaches for analysis of deep-phenotypic psychiatric data in neurogenetic disorders.

## INTRODUCTION

Genetically defined disorders that impact brain development are not only important medical conditions in their own right, but also provide naturally-occurring opportunities to study the consequences of high genetic risk in humans more generally (1–3). Recurrent gene dosage disorders (GDDs) - including sub-chromosomal copy number variations (such as deletions or duplications at 22q11.2 or 16p11.2) and aneuploidies of chromosomes X, Y or 21 (2) - represent a major class of genetic risk for neuropsychiatric illness. The recurrent nature of these disorders makes it possible to recruit and characterize groups of individuals with the same gene dosage abnormality (3). Phenotypic characterization of GDDs at neuroanatomical and transcriptomic levels has benefitted from access to high-dimensional data (e.g. measures of many brain regions (4–6) or genes (7)) and advanced analytic approaches (e.g. network-based modeling of brain regions (5) or genes (7)), but there has been limited application of such research strategies to understand symptomatology in GDDs. This disparity in research approaches is problematic given that behavioral manifestations of GDDs are the immediate concern of carriers, caregivers, and clinicians. More fully understanding the complex behavioral manifestations of GDDs is not only clinically relevant but arguably necessary for meaningful research on biological factors that may predict clinical outcomes. This need is all the more important given the rapid growth and strategic prioritization of research on rare genetic disorders that increase risk for psychopathology (2).

Here, we present research approaches for bringing high-dimensional phenotypic and network-analytic methods to bear on behavioral characterization of GDDs. We illustrate these generalizable approaches through initial application to XYY syndrome. XYY syndrome is a historically stigmatized sex chromosome aneuploidy (8–10) that remains poorly characterized despite affecting 1 out of every 1000 males (11,12), substantially increasing risk for a range of behavioral and emotional difficulties (13,14), and providing a naturally occurring model for Y-chromosome effects on the human brain (15,16). To date, the nature of psychiatric risk in XYY syndrome has never been established using gold-standard interviewer-based diagnostic instruments. Moreover - as for most GDDs in psychiatry - few studies of XYY syndrome have considered diverse psychiatric domains in parallel (17,18) to (i) pursue a comprehensive mapping of psychopathology, (ii) uncover the relationships between behavioral concerns in different domains, and (iii) discern how different aspects of psychopathology relate to co-occurring variation in cognitive ability, adaptive functioning and caregiver strain. We sought to develop approaches for addressing these general issues in GDD research, and the current study applies this framework to build on pioneering earlier XYY studies (17,18) in three key directions.

We first assess psychiatric morbidity in youth with XYY syndrome at the diagnostic-level using the K-SADS (Kiddie Schedule for Affective Disorders and Schizophrenia) (19) – an interviewer-based gold-standard diagnostic instrument that involves structured interviews with affected individuals as well as their primary caregivers. Within a cohort of 64 individuals with XYY syndrome, we use the K-SADS to first estimate psychiatric disorder prevalence rates using both full diagnostic and screening criteria, and then to quantify rates of diagnostic comorbidity and their relationships with global measures of cognitive ability, adaptive function, and caregiver strain. We also integrate categorical diagnostic ratings from the K-SADS with dimensional measures of psychopathology from the Child Behavior Checklist (CBCL) (20) in order to characterize symptom burden outside of formal diagnostic thresholds and to assess the sensitivity and specificity of CBCL cut-offs for K-SADS diagnoses. These latter two goals directly inform clinical practice in assessment of XYY individuals by clarifying the degree to which diagnostic status adequately captures observed levels and patterns of psychopathology in XYY syndrome, and the degree to which questionnaire-based measures can be used as a screen for diagnostic status.

Next, we systematically map the patterned effect of XYY syndrome across 67 dimensional measures of psychiatric symptomatology from 12 different rating scales. This multidimensional measurement strategy provides intentionally redundant estimates of symptom severity from distinct tools to characterize diverse domains spanning attention, impulse control, social reciprocity, mood, repetitive behaviors, motor coordination, and aggression. Considering so many domains in parallel allows for a more fine-grained understanding of strengths and difficulties in XYY syndrome. We model these high-dimensional behavioral data by borrowing analytic methods developed in network science (21) and recently applied in neuroscience (22) to build network-based representations of symptom variation in XYY syndrome, which we then use to (i) identify a reduced set of symptom clusters that underlie the diversity of behavioral presentations across individuals with XYY syndrome, and (ii) nominate clinically salient behavioral features that may be especially closely tied to IQ, adaptive functioning and caregiver strain.

Finally – for both diagnostic and dimensional outcomes in XYY - we compare prenatally vs. postnatally diagnosed XYY subgroups as a means (23) of probing potential genetic diagnosis ascertainment biases that can complicate the study of gene dosage disorders. Although postnatal diagnosis of XYY syndrome is often precipitated by concerns regarding behavioral and developmental difficulties (24), this source of ascertainment bias cannot apply to prenatally diagnosed individuals. Thus, phenotypic profiles in prenatally diagnosed individuals may more accurately estimate the true penetrance of XYY and better generalize to the large fraction of XYY individuals who remain undiagnosed (24). Moreover, the phenotypic profile of prenatally diagnosed individuals with XYY syndrome is likely to become more clinically representative with increasing access to prenatal genetic testing (25).

## METHODS

### Participants

Our dataset included a total of 124 individuals (**Table 1**). Sixty-four participants with XYY syndrome (ranging in age from 5 to 25 years; mean age 13.1 years, SD 5.7) were recruited with the help of the Association for X and Y Chromosome Variations (AXYS; genetic.org) and the National Institutes of Health (NIH) Clinical Center Office of Patient Recruitment. We also recruited a cohort of 60 age- and sex-matched typically developing controls, who were used to derive normative reference scores for those measurement scales that did not provide scaled scores relative to their own population norms (for details, see below). All controls were screened using a standardized interview to verify the absence of a prior psychiatric diagnosis or any early developmental difficulty requiring provision of extra support at school or home. The sole inclusion criteria for XYY individuals in this study were cytogenetically confirmed XYY karyotype and age between 5 and 25 years on the day of assessment. For those XYY participants who were able to give blood (*n*=37), the presence of XYY was re-confirmed through repeat karyotype testing based on observation of non-mosaic XYY karyotype across a minimum of 50 metaphases (Quest Diagnostics, Nichols Institute Chantilly, VA). For the remaining participants, the presence of non-mosaic XYY karyotype was verified by inspection of existing community-based genetic testing reports. Exclusion criteria shared by both XYY participants and controls were a history of brain injury or comorbid neurological disorders. All research assessments were conducted at the NIH Clinical Center, Bethesda, MD, USA. Written informed consent was secured from adult participants and parents of minor participants and written assent was secured from all children. The National Institute of Mental Health (NIMH) Institutional Review Board approved the study.

**Table 1:**
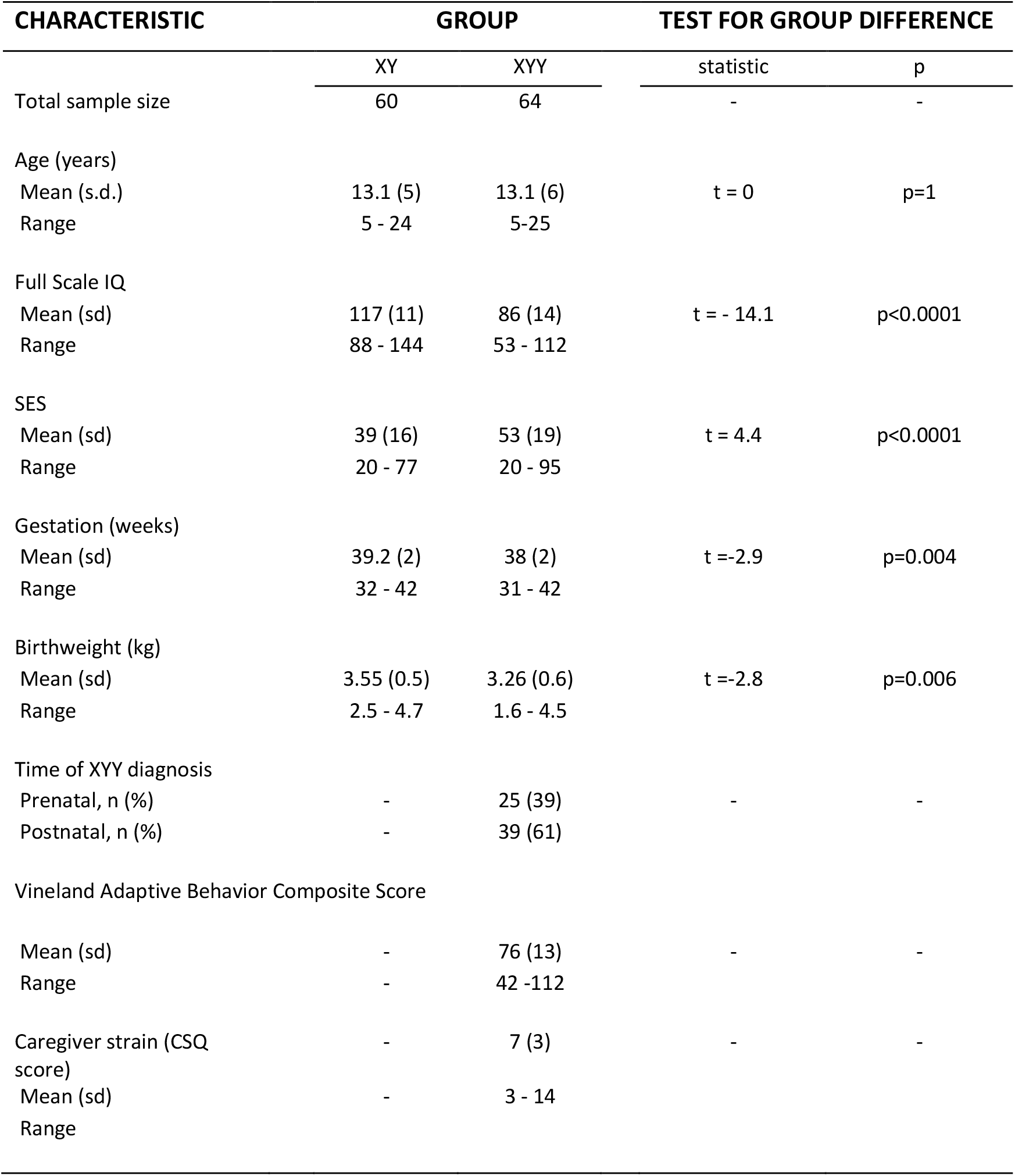
Participant Characteristics.

### Participant Characterization

#### Clinical History

All XYY participants received a structured medical history and physical examination. Caregiver reports and reviews of prior medical documents were used to record the age at which XYY was diagnosed. Caregiver strain was measured using the Caregiver Strain Questionnaire [CSQ, (26)]

#### Diagnostic Assessments

The K-SADS (19) was administered to all XYY participants by a trained psychiatric nurse practitioner (ET), and all final consensus ratings were reviewed with a child and adolescent psychiatrist (AR). The K-SADS is a semi-structured instrument used with both the participant and parent/guardian assessed separately to diagnose and screen for various psychiatric disorders based on the Diagnostic and Statistical Manual of Mental Disorders-Fifth Edition (DSM-5) guidelines. The K-SADS was used to generate diagnostic prevalence rates using standard DSM-5 criteria (henceforth referred to as “full” diagnostic criteria). We also generated “sub-threshold” prevalence rates for each disorder based on the number of individuals who endorsed the K-SADS screening questions for that disorder (henceforth referred to as “screening diagnostic criteria”), regardless of whether they met full diagnostic criteria based on relevant post-screen K-SADS supplement questions. The K-SADS was also used to gather information regarding developmental milestones as well as educational, medical and prenatal history. All components of the K-SADS were administered in this study, with the exception of the Autism Spectrum Disorder (ASD) section. All XYY participants underwent a formal research assessment of ASD features based on an ASD diagnostic battery with three components: the Autism Diagnostic Observation Schedule, 2^nd^ Edition [<u>ADOS; 21</u>], the ADI-R (27), and the DSM-5 diagnostic criteria checklist [<u>22</u>]. Measures were performed by licensed clinical psychologists (L.J., A.T., C.C.) with extensive ASD evaluation experience, who met research reliability standards on the ADI-R and ADOS-2. For descriptive purposes, we also calculated collective diagnostic rates for groups of disorders defined as follows (**Table 2**): Neurodevelopmental Disorders [including ASD, Attention Deficit Hyperactivity Disorder (ADHD), Tic Disorder, Obsessive Compulsive Disorder]; Mood Disorders (including Bipolar Disorder, Depressive Disorder, Disruptive Mood Dysregulation Disorder); Anxiety Disorders; Posttraumatic Stress and Related Disorders; Disruptive, Impulse Control and Conduct Disorders (including Oppositional Defiant Disorder and Conduct Disorder); and Substance Use Disorders.

**Table 2.**
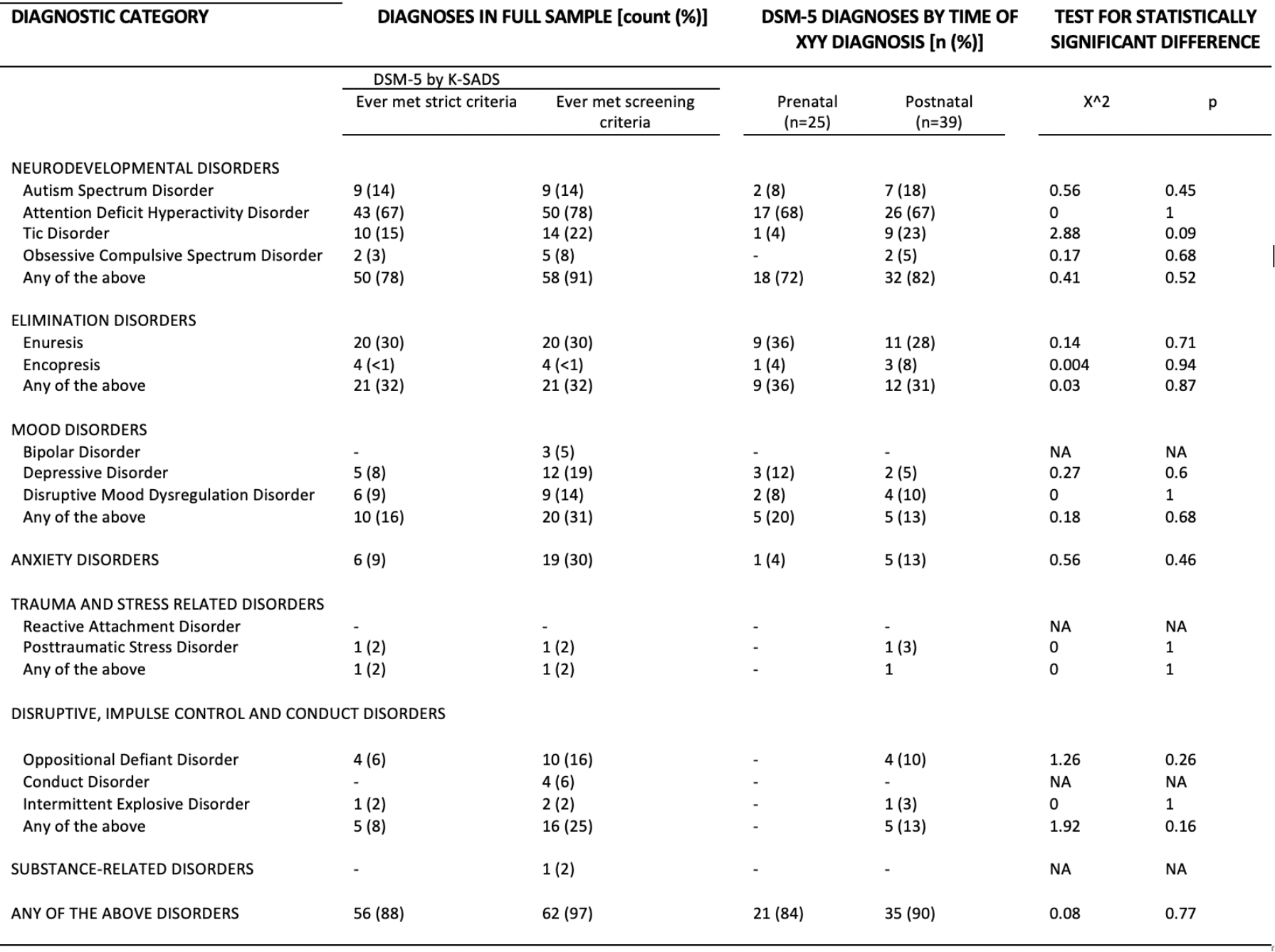
Psychiatric Diagnostic Rates in the XYY Syndrome. Diagnostic rates based on the K-SADS (and ADI/ADOS for ASD) are provided using strict and screening criteria (where screening criteria refers to the endorsement of at least one K-SADS screening question). We also show diagnostic rates stratified by the time of XYY diagnosis, and tests for the statistical significance of differences in diagnostic rates between pre- and postnatally diagnosed subgroups.

#### Child/Adult Behavior Checklist (referred to collectively as CBCL

The CBCL is an extensively used and norm-referenced set of questionnaires for dimensional measurement of psychopathology (28). Individual items in the CBCL are summed to provide dimensional measures of eight independent empirically-derived subscales (referred to as “syndrome subscales” by CBCL developers: Anxious/Depressed, Withdrawn/Depressed, Somatic Complaints, Social Problems, Thought Problems, Attention Problems, Rule Breaking Behavior, and Aggressive Behavior), two composite scales combining information across some of these subscales (Internalizing and Externalizing Problems), and a single summary measure of Total Problems (**Table 3**). This instrument has also been used in several independent natural history studies of XYY and other genetically-defined neurodevelopmental syndromes (18,29) – facilitating direct comparison with our findings in this new deeply phenotyped cohort.

**Table 3.**
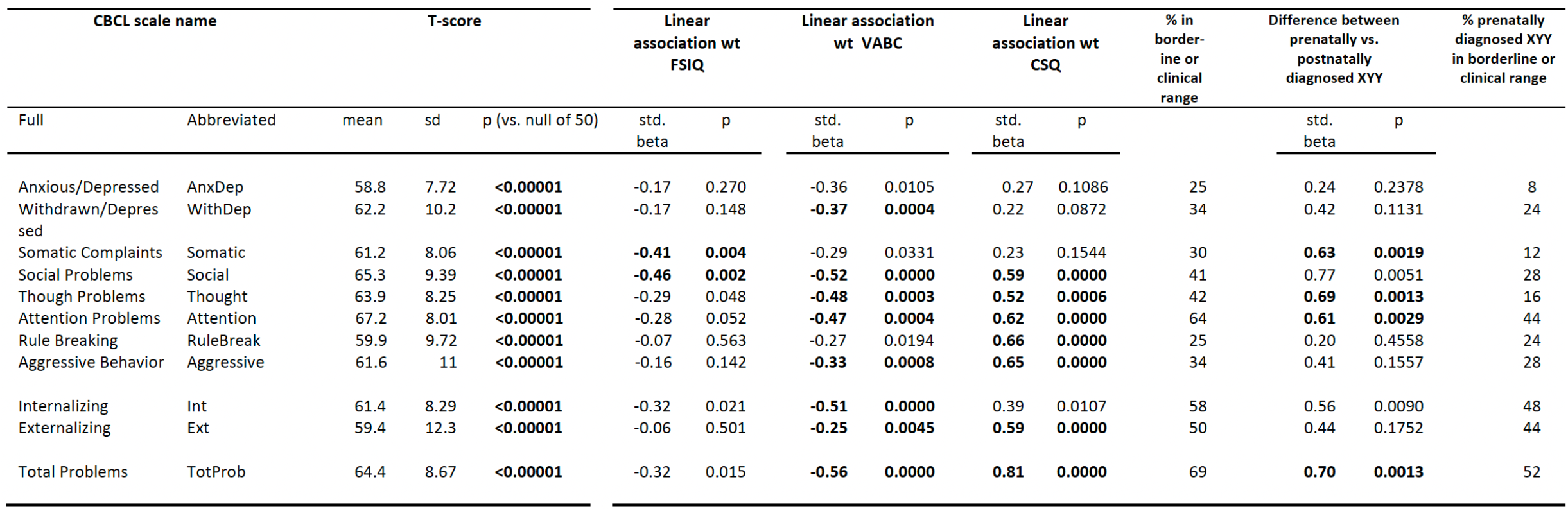
Child Behavior Checklist Scores in the XYY Syndrome. Score distributions are given for each subscale, both broadband scales and the Total Problems scale. We also show the standardized regression coefficient and associated *p*-values for regressions using interindividual score variation in each CBCL scale to model variation in Full Scale IQ (FSIQ), Vineland Adaptive Behavior Composite Scale (VABC) and the caregiver strain questionnaire (CSQ). Finally, we report: the percentage of all 64 individuals with CBCL scale scores in the borderline or clinical range; the regression effect size and *p-*value for prenatally vs. postnatally diagnosed subgroups of the full XYY cohort; and the percentage of all 25 prenatally diagnoses XYY individuals with CBCL scale scores in the borderline or clinical range. Bold text denotes statistical tests that survive Bonferroni adjustment for multiple comparisons (adjusted *p-*value=0.05/11=0.0045).

#### Full Battery of Questionnaire-Based Measures of Behavior and Psychopathology

To provide a broader and deeper phenotypic assessment, we also gathered information using several diverse questionnaire-based measures other than the CBCL. These instruments provided finer-grained information regarding several dimensions of psychopathology including features of autism spectrum disorder (e.g. Social Responsiveness Scale, SRS, (30)), obsessive-compulsive disorder (e.g. The Obsessive-Compulsive Inventory, OCI-R, (32)), motor coordination disorder (e.g. Developmental Coordination Disorder Questionnaire, DCDQ, (33)), attention deficit hyperactivity disorder (e.g., Conners-3, (34)), impulsivity (e.g. Barratt impulsiveness scale, BIS, (35)), conduct/dissocial disorders (e.g. antisocial process screening device, APSD, (36)), and aggression (e.g. Children’s Scale of Hostility and Aggression, C-SHARP, (37)). We also used the Strength and Difficulties Questionnaire (SDQ, (38)) as a multi-dimensional measure of childhood psychopathology complementary to the CBCL. Thus, our full battery of questionnaire-based measures (full annotated list in **Table 4**) provided a set of 67 continuous variables (the number of sub-scales across instruments) which collectively spanned most major domains of psychopathology in youth. The redundancy between sub-scales in the battery was an intentional aspect of our study design as it allows for the same construct to be captured by different instruments and provides a means of assessing whether observed cross-trait correlations are organized according to shared phenomenology as opposed to more superficial features such as instrument of origin.

#### Cognitive Assessments

The Wechsler Preschool and Primary Scale of Intelligence, Fourth Edition, Wechsler Intelligence Scale for Children, Fifth Edition, or Wechsler Adult Intelligence Scale, Fourth Edition was used to assess intelligence. If the participant had been tested with a Wechsler scale within 1 year (n=4), the Wechsler Abbreviated Scale of Intelligence, Second Edition was used. We used these instruments to generate a single Full Scale Intelligence Quotient (henceforth “IQ”) score for each study participant.

### Statistical Analysis

Categorical variables were described using counts and rates, and continuous variables using means and standard deviations. Chi-squared tests and t-tests were used to compare categorical and continuous (respectively) variables as a function of diagnostic status for individual DSM-5 categories (present vs. absent).

#### Diagnostic Categories

Chi-squared tests were used to compare diagnostic rates as a function of age at XYY diagnosis (prenatal vs. postnatal). Linear regression was used to relate the total number of diagnoses to three global measures of cognitive ability and functioning: IQ (as measured by Weschler intelligence scales, see above), adaptive functioning (the Vineland Adaptive Behavior Scale, 2^nd^ edition (VABC-II) adaptive behavior composite score, VABC (39)) and caregiver strain total score (Caregiver Strain Questionnaire, CSQ, (26)).

#### Child Behavior Checklist

One tailed t-tests were used to determine which CBCL scales had score distributions in XYY that were significantly shifted relative to population norms (i.e., scales for which the estimated mean for the percentile score distribution in XYY was statistically significantly different from 50). The distribution of CBCL scores for each domain was visualized by plotting t-scores rather than percentile scores. Although CBCL *t*-scores are strongly correlated with percentile scores, they can be used to identify people who fall into a “clinical risk” category for each scale (i.e. *t*-scores > 65 for subscales and >60 for “broad band” scales). The CBCL *t*-scores were also used to examine the interrelationship between diagnostic and dimensional measures of psychopathology in XYY syndrome as follows: to characterize clinical risk in those falling below diagnostic thresholds we calculated - for each K-SADS diagnosis and CBCL scale pair - the proportion of individuals without the K-SADS diagnosis who had CBCL scale *t*-scores in the clinical risk category. We also quantified the sensitivity and specify of CBCL clinical risk category for each K-SADS diagnosis. Pairwise relationships between CBCL clinical risk status and K-SADS diagnostic status were visualized as heatmaps. All one-sample t-tests of CBCL *t*-score distributions and linear regression models of t-score variation were adjusted for multiple comparisons using Bonferroni correction (adjusted p value=0.05/11=0.0045).

#### Full Battery of Questionnaire-Based Measures of Behavior and Psychopathology

To profile behavior and psychopathology in XYY across our full battery in a manner that allowed comparison between different instruments, we expressed all 67 sub-scale scores in XYY as z-scores relative to the distribution observed in our independent recruited sample of typically developing male controls (**Table S1**). Since the instruments used did not consistently provide normed scores, we used the following procedure to bring all XYY sub-scale scores into a common reference frame relative to score distributions in the 60 age matched XY controls. We first tested for the presence of a statistically significant difference in age effects on scale scores (scaled scores if available and raw if not) between XYY case and XY controls. Where such age-by-group interactions were found, we re-expressed observed scores for all XYY and XY individuals as standardized residuals from predicted scores for their age given by a general linear model for score as a function of age estimated in our XY control cohort. In the absence of such age-by-group interactions, we expressed XYY scores for a scale as z-scores using the distribution in XY controls. In this framework, all group differences in scale scores represented standardized effect size shifts of XYY relative to XY controls. Statistically significant differences in scaled scores between XYY and XY groups were identified using linear models with scale score as a continuous dependent variable and group as a categorical predictor – for the full XYY group as well as the subgroup of prenatally diagnosed individuals. We verified that findings from these linear models agreed with those from non-parametric Wilcox Rank Sum tests. For each scale, we also computed standardized regression coefficients for the relationship between score variation in the XYY group and IQ, VABC, CSQ and timing of XYY diagnosis (prenatal vs. postnatal). All statistical tests were Bonferroni-adjusted for multiple comparisons across scales (adjusted p value = 0.05/67=0.0007).

#### XYY Behavioral Network Analyses

For the XYY group, we also modelled inter-relationships among all dimensional measures in our battery – using scores that had been scaled relative to the XY group as described above. For these network analyses, we excluded (i) any measures that were available for less than 2/3 of XYY participants (BIS, DASS and OCI), and (ii) any subscales with low overall network connectivity - defined as a mean weighted degree below the 5^th^ centile of the distribution from all scales (Conners – learning subscale, SCQ-repetitive behavior subscale, CBCL – withdrawn/depressed subscale) leaving a total of 63 behavioral scales for downstream analysis (**Table S1**). We generated a square matrix of pairwise Pearson correlations between these 63 measures. This matrix can be conceptualized as an unthresholded weighted network where behavioral measures are nodes and correlation coefficients define the edge strength (weight) between each pair of nodes. We clustered this matrix using weighted stochastic block modeling (WSBM) as implemented using the *BM_gaussian* function with default setting from the R package *blockmodels* (40). We chose the WSBM for its generalizability to the problem of detecting clusters of various sorts in complex networks. Typical methods for cluster detection tend to identify clusters comprised of nodes that are highly similar to each other and highly dissimilar to other nodes; such clusters are called assortative, as they contain nodes of similar connectivity profiles. The WSBM, by contrast, is a generative modelling technique that is capable of detecting non-assortative clusters, which contain nodes of dissimilar connectivity profiles (22). An intuitive example of a non-assortative clusters is a “core-periphery” structure, wherein a core of densely interconnected nodes extends projections to a periphery of sparsely interconnected nodes. In the context of behavioral scales this core might represent a general psychopathology cluster for example, with peripheral clusters representing distinct sub-components of psychopathology. In application to our dataset, the WSBM algorithm assigned the 63 behavioral nodes into a smaller number of clusters and estimated a mean correlation between each cluster pair. We used this lower-dimensional representation of behavioral variation in XYY syndrome to: (i) generate a network visualization capturing the sub-structure of behavioral variation in XYY syndrome, and (ii) calculate person-specific z-scores for each cluster representing the average of z-scores for all nodes/scales in the cluster (see above for details of z-score generation for each scale in the XYY group), which could then be related to individual variation in the global cognitive and functional measures of IQ, VABC and CSQ.

## RESULTS

### Participant Characteristics

Our study included 64 singletons with XYY syndrome aged between 5 and 25 years (**Table 1**), and 60 age- and sex-matched controls who provided reference distributions for those measures of psychopathology and behavior that lacked published norms. Relative to XY controls, XYY participants had a significantly lower mean IQ, socioeconomic status (SES, (41)), gestational age and birthweight. The majority of XYY participants (*n*=39, 61%) received their genetic diagnosis after birth (mean age of diagnosis=6 years, range 0.02 years to 16 years), and - as previously reported (23) - the mean full scale IQ of this postnatally diagnosed group was lower than that of the XYY subgroup who had been diagnosed prenatally or at birth (FSIQ 83 vs. 90, t=2, p=0.05).

### Psychiatric Diagnoses in XYY Syndrome

According to K-SADS and ASD diagnostic assessment, the overall prevalence rate for ever having had any DSM-5 psychiatric diagnosis by the time of assessment was 82% in the full sample of XYY individuals (**Table 2**). Neurodevelopmental disorders were the most prevalent diagnostic group (prevalence of ever having met diagnostic criteria = 78%), followed by elimination disorders (32%), mood disorders (16%), anxiety disorders (9%) and disruptive, impulse control and conduct disorders (8%). The three most prevalent diagnoses were Attention Deficit and Hyperactivity Disorder (ADHD, 67%), Enuresis (30%), and ASD (14%). We observed high rates of comorbidity amongst DSM-5 diagnoses in XYY syndrome (**Fig. 1A**), with a modal number of diagnoses of 2 (range 0-5). Over half (55%) of participants had met criteria for two or more psychiatric diagnoses by the time of assessment. Greater diagnostic comorbidity was associated with significantly lower IQ and adaptive functioning, as well as higher caregiver strain (**Fig. 1B-D**). As expected, use of K-SADS screening (as opposed to full) criteria led to “screening diagnosis” rates that were higher that strict diagnostic criteria (e.g. 96% of individuals met screening criteria for one or more disorders by the time of their participation in the study) and also elevated diagnostic comorbidity (modal number of conditions=3, maximum=7) (**Table 2, Fig. 1A**). Finally, rates of psychiatric diagnosis tended to lower amongst the prenatally diagnosed subgroup of XYY individuals relative to the postnatally diagnosed subgroup, although these differences did not reach statistical significance (**Table 2**). Seventy-six percent of participants in the prenatally diagnosed XYY subgroup had met screening criteria for one or more psychiatric diagnoses by the time of entry into the study.

**Figure 1.**
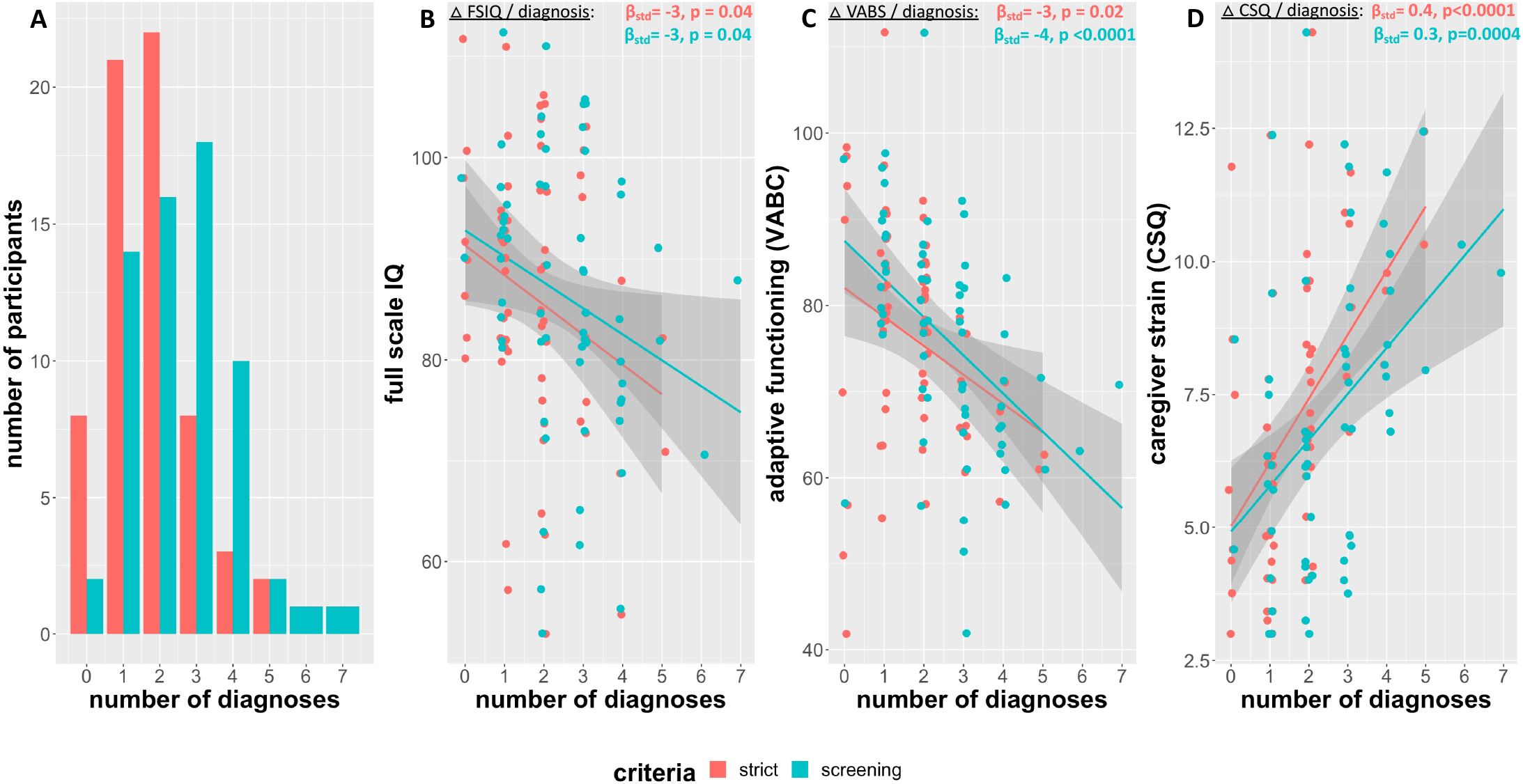
Diagnostic Comorbidity in XYY Syndrome. **A**. Histogram showing participant distribution by number of diagnoses for both strict (red) and screening (blue) diagnostic thresholds. B, C, D. Scatterplot of relationship between number of diagnoses and Full-Scale IQ (**B**), Vineland Adaptive Behavior Composite Score (VABC) (**C**) and Caregiver Strain Questionnaire (CSQ) score (**D**).

### Dimensional Measures of Psychopathology in XYY Syndrome from the CBCL

The CBCL revealed a patterned elevation across multiple domains of psychopathology in XYY syndrome, with lowest mean subscale scores in Anxious/Depressed and Rule Breaking, and highest in Attention and Social Problems (**Table 3, Fig. 2A)**. We observed a partly subscale-specific relationship between participant CBCL scores and participant IQ, participant adaptive functioning and caregiver strain (**Table 3**). For example, the CBCL Externalizing scale was uncorrelated with FSIQ but showed the strongest positive association with caregiver strain (Pearson *r* > 0.7). However, the CBCL Social Problems subscale was unique in showing a significant association with all three of these outcomes and was the single most predictive CBCL subscale for FSIQ and adaptive functioning (all Pearson *r* < −0.4). Those CBCL domains with more severe disruption in XYY syndrome were also those for which interindividual score variation was most strongly related to variation in FSIQ and adaptive behavior (**Fig. 2B**) – suggesting a close relationship of social and attentional impairments in XYY syndrome with general cognitive and adaptive functioning.

**Figure 2.**
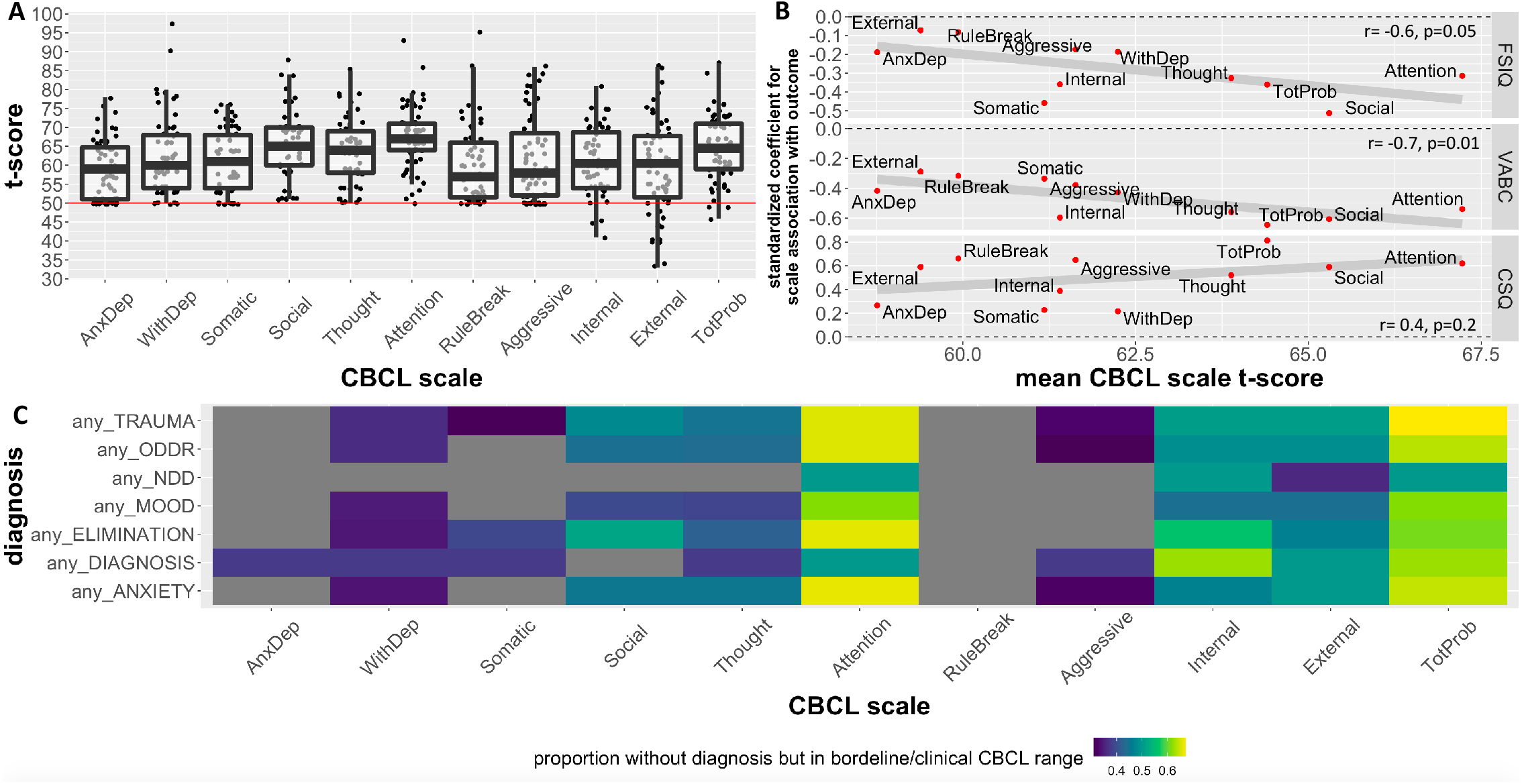
Childhood Behavior Checklist Measures of Psychopathology in XYY syndrome. **A**. Dot and boxplot showing distribution of-scores for CBCL sub-scales and broadband scales in XYY syndrome. The normed average of 50 is shown as a horizontal red line. **B**. Dot and line plot showing relationship between each CBCL scale’s mean t-score, and the degree to which variation in scale scores is related to variation in IQ, adaptive function [Vineland Adaptive Behavior Composite Score (VABC)] and caregiver strain [Caregiver Strain Questionnaire (CSQ)]. **C**. Heatmap showing the proportion of XYY individuals lacking each diagnosis (rows) that have CBCL scores in the clinical risk range – for each CBCL scale (columns).

CBCL subscale *t*-scores higher than 65 (and broad-band t-scores >60) are taken to indicate that an individual is in the borderline or clinical range. Based on CBCL norms, less than 3% of the general population would score above this t-score cut-off for any given CBCL scale. For most CBCL subscales – over 30% of the XYY participant had scores within the borderline or clinical range (**Table 3**). Examining CBCL borderline/clinical status as a function of the presence versus absence of psychiatric diagnoses revealed that for many psychiatric disorder categories, a significant proportion of XYY participants who fell below DSM-5 diagnostic cut-offs still achieved borderline or clinical range CBCL scores (**Fig. 2C**).

CBCL scores tended to be less elevated in the prenatally vs. postnatally diagnosed group, with statistically significant subgroup differences for CBCL Total, Internalizing, Thought, Somatic and Attention scales (**Table 3**). Nevertheless, borderline or clinical range CBCL scores were still seen for over 20% of participants in the prenatally diagnosed subgroup for Total, Externalizing, Attention, Aggressive, Rule Breaking, Anxious/Depressed and Social Problems scales (**Table 3**). Sixty-four percent of prenatally diagnosed individuals fell outside the borderline or clinical range for one or more CBCL subscales.

### Score Profiles in XYY Syndrome Across a Large Battery of Questionnaire-Based Measures

The 12 questionnaires within our full battery collectively provided a total of 67 subscale and summary scale scores which allowed us to develop fine-grained maps of behavior and psychopathology in XYY syndrome. The number of participants with available scores for a given scale/subscale ranged between 25 and 62 due to differences in the applicable age-range for each questionnaire. However, the XYY and XY groups with available scores for each scale/subscale were similarly sized and did not differ in mean age (**Table S1**).

We first expressed score distributions in the XYY group as z-score relative to score distributions observed in our sample of XY controls. This procedure allowed us to rank all 67 measured dimensions by their relative disruption (median scaled score) in XYY syndrome in a single combined boxplot visualization (**Fig. 3A**), and to determine the magnitude and statistical significance of score distribution differences between XYY and XY groups (**Table S1, Fig. 3B**). The resulting ranking of scales was broadly organized by psychopathological domain rather than questionnaire of origin: the greatest score elevations were seen in questionnaire subscales that indexed global social functioning (e.g. standardized β ∼ 6.5 for the CBCL social difficulties subscale); the smallest score elevations were seen in subscales relating to obsessive, aggressive and dissocial behaviors (e.g. standardized β = 0.6 for the OCI-R Obsessions scale); and intermediate elevations were seen for subscales capturing cognitive, motor, attentional and mood difficulties. After correction for multiple comparisons, 60 (83%) of measured dimensions showed statistically significant score elevations in the XYY vs. XY group (**Table S1**). The few subscales that did not show statistically significant score elevation in XYY as compared to XY groups mostly related to obsessive-compulsive behaviors and physical aggression.

**Figure 3.**
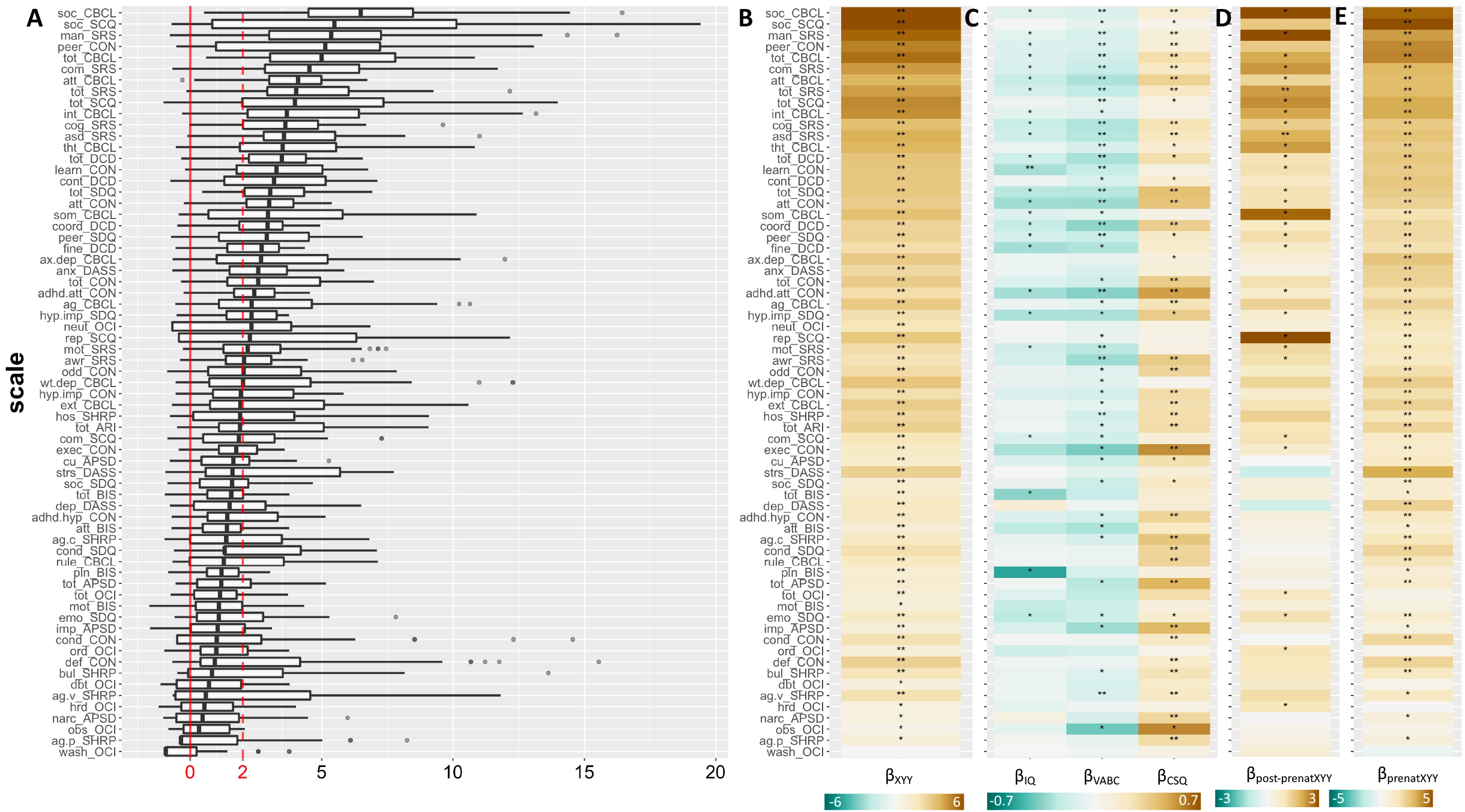
Profiling Symptoms in XYY Syndrome Across 67 Scales. **A**. Barplots showing distribution of scaled scores in XYY syndrome for 67 different subscales derived from 10 questionnaires. Vertical solid red line shows mean score in XY controls, and dashed vertical line shows a 2 standard deviation elevation in scores relative to the observed distribution in XY males. **B**. Standardized beta coefficients for the effects of XYY (vs. XY) on each scale. **C**. Standardized beta coefficients for regression models predicting variation in IQ (Full Scale IQ), VABC (Vineland Adaptive Behavior Composite Scale) and CSQ (Caregiver Strain Questionnaire) scores as a function of variation in each measure of psychopathology. **D**. Standardized beta coefficients for association between time of XYY diagnoses (postnatal vs. prenatal) and scores on each measure of psychopathology. **E**. Standardized beta coefficients for the effects of XYY (vs. XY) on each scale when analysis is restricted to the subset of XYY Individuals who were diagnosed prenatally. For panels (B-E), a single asterisk indicates statistical significance at uncorrected p=0.05, and a double asterisk at false discovery rate (FDR) adjusted *q*=0.05.

We observed a scale-specific relationship between variation in symptom severity and variation in participant IQ, participant adaptive behavior and caregiver strain (**Table S1, Fig. 3B**). Patterned associations with psychopathology were strongest for CSQ, then VABC then IQ (mean standardized β across scales =0.22, −0.15 and −0.1 respectively). After correction for multiple comparisons; variation in IQ was significantly associated with the Conners Learning Problems scale (standardized β = −0.5) and variation in VABC and CSQ were both associated with variation in multiple scales (n=22 and n=35, respectively) with shared associations for social and attentional impairments. Notable dissociations included a unique association of VABC with measures of coordination and learning problems (total DCD score and Conners learning subscale), and a unique association of CSQ with multiple scales capturing disruptive and externalizing behaviors. Scale scores tended to show greater elevation in postnatally vs. prenatally diagnosed XYY subgroups (**Table S1, Fig. 3D**, mean standardized β for time of XYY diagnosis on scale score = 1, one sample *t-*test vs. 0: t= 10.5, p= 1.2*10^−15^). After correction for multiple comparisons, these subgroup differences only reached statistical significance for 2 ASD-related measures (SRS-2 Total and DSM-compatible ASD Social Communication/Interaction scale scores). Accordingly, compared to XY controls, the subgroup of prenatally diagnosed XYY individuals still showed statistically significant score elevations in 53 (76%) of the dimensions examined. Moreover, the profile of relative impairment across subscales in the prenatally diagnosed XYY subgroup relative to XY controls was highly similar to that for the XYY group as a whole (Pearson correlation of standardized β across scales *r* > 0.9).

### Identifying Core Components of Behavioral and Psychiatric Difficulties in XYY Syndrome

Weighted Stochastic Block Modelling (WSBM, **Methods**) of the inter-scale correlation matrix grouped scales into 8 clusters which each tended to combine scales that came from different instruments but were often related to a similar theme. We named these 8 clusters as follows based on their scale content (presented in descending order of mean cross-cluster correlation): Total Psychopathology, Inattention, Externalizing, Current Social Impairments, Impulsivity, Internalizing, Dissociality and Early Social Impairments. Heatmap (**Fig. 4A**) and graph-based (**Fig. 4B**) visualization of this WSBM solution revealed several features of note. Total Psychopathology occupied a central hub-like position in the psychopathology network and showed strongest connectivity with a tightly interrelated set of four clusters: Inattention, Externalizing, Impulsivity and Current Social Impairments. Thus, these five behavioral clusters appear to form a core element of the XYY psychopathology network, with other clusters occupying more peripheral network positions that broadly delineate three relatively weakly inter-related aspects of psychopathology in XYY syndrome: Internalizing problems, Dissociality and Early Social Impairments. Cluster degree within the network showed a moderate positive correlation (r=0.4) with the mean z-score of scales within the cluster (i.e. the average across all cluster scales and all individuals of participant-level z-scores generated for XYY participants as described above). Finally, we generated person-level mean z-scores for each cluster (i.e. the within-person, cross-scale average of scale-specific z-scores generated as described above), and estimated the relationship between interindividual variation in cluster scores and variation in IQ, VABC, and CSQ. Echoing our findings at the level of individual subscales, we found that the cluster scores showed variable patterns of statistically significant correlations with IQ, VABC, and CSQ. For example, the Impulsivity and Externalizing clusters were significantly correlated with CSQ alone; Total Psychopathology was significantly correlated with VABC and CSQ; whereas Inattention and Current Social Impairments were significantly correlated with all three measures.

**Figure 4.**
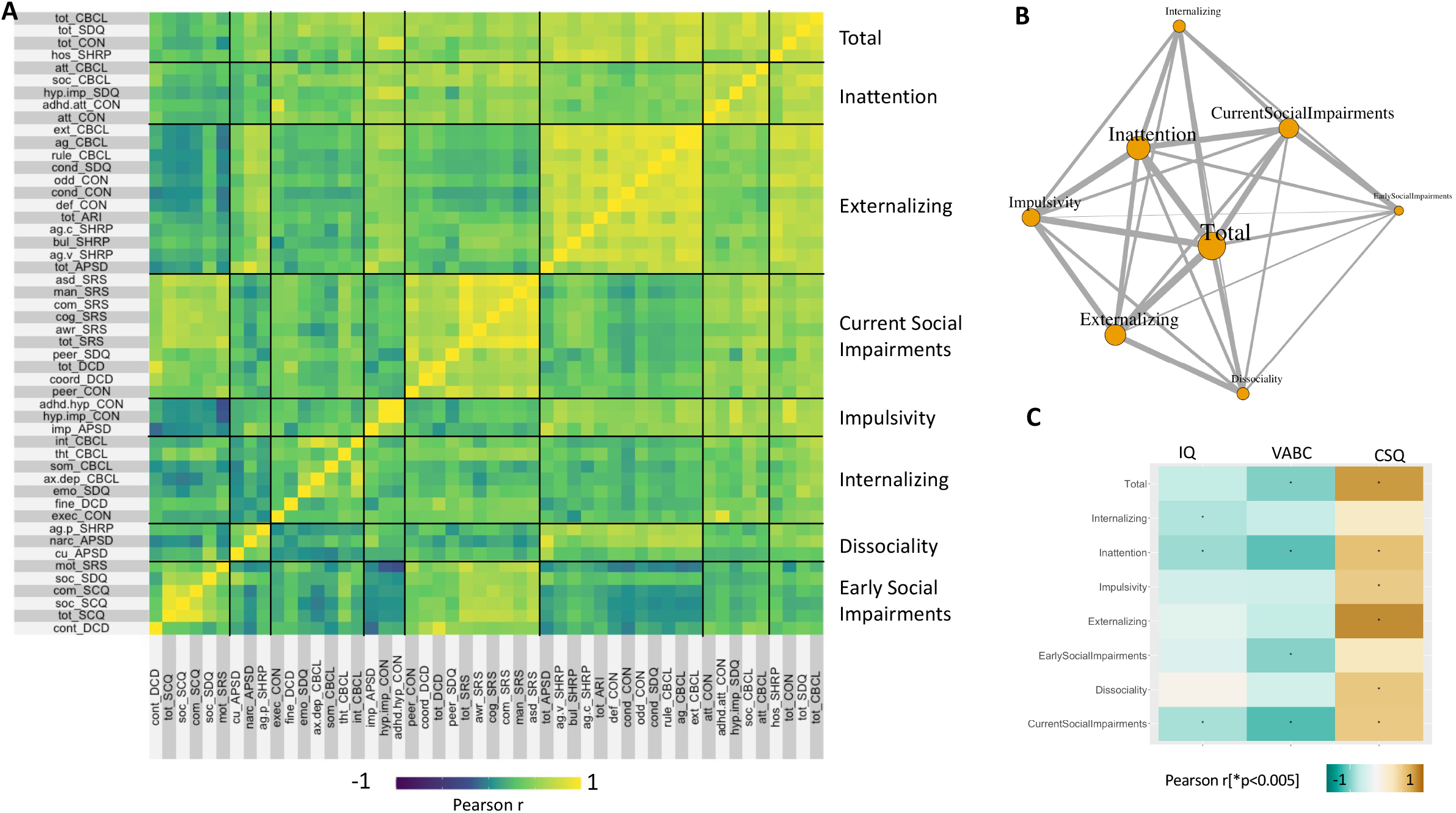
Network Architecture of Psychopathology in XYY Syndrome. **A**. Heatmap showing Pearson correlation coefficients between pairs of scales across individuals with XYY syndrome. Scales are grouped according to the optimum *k*=8 Weighted Stochastic Block Model (WSBM) solution, and black lines define boundaries between blocks defining pairwise correlations between measures within and between each cluster. **B**. Network representation of the WSBM solution shown in panel (A). Each node is one of the 8 clusters in panel (A). Edges show the WSBM estimated mean correlation between scales in each cluster (edge width indexes correlation magnitude: thicker indicates more positive). Node size represents weighted degree based on these edge strengths. Graph layout is with the Fruchterman-Reingold force-directed algorithm. **C**. Heatmap showing correlations (Pearson *r*) between person-level WSBM cluster scores and variation in IQ (Full Scale IQ), VABC (Vineland Adaptive Behavior Composite Scale) and CSQ (Caregiver Strain Questionnaire) scores. Asterisk indicates statistical significance at *p*<0.005.

## DISCUSSION

The analytic approaches implemented in this study provide several novel insights into the psychiatric manifestations of XYY syndrome, and thereby illustrate the potential value of extending such methods to the many other complex neurogenetic disorders that are of clinical and research interest in psychiatry.

First, we estimate the prevalence of psychiatric diagnoses in XYY through extensive in-person interviews using gold-standard diagnostic instruments, and we complement this diagnostic view with dimensional measures of psychopathology. We find that carriage of an extra Y-chromosome can have substantial negative impacts on several mental health outcomes – even in subgroups with reduced genetic diagnosis ascertainment bias by virtue of having been diagnosed prenatally. Notwithstanding the high variability in outcomes across individuals, most individuals in the full XYY cohort met criteria for at least one psychiatric diagnosis by the time of entry into our study – and this proportion remained high (>75%) for those who received their XYY diagnosis prenatally. Diagnostic rates of ASD and ADHD showed an approximately four-fold elevation above those in males from the general population (42,43), and we also observe elevated risk for many other neurodevelopmental, mood and anxiety disorder diagnoses. The majority of XYY individuals had met criteria for more than one psychiatric diagnosis by the time of enrollment, and greater diagnostic co-morbidity was significantly associated with lower IQ, worse adaptive functioning, and greater caregiver strain. Combined analyses of diagnostic and CBCL data reveals statistically significant elevations across all CBCL domains in the full XYY group, and substantial psychiatric morbidity even amongst participants that fall below diagnostic thresholds. The CBCL subscales most impacted by XYY syndrome (Attention and Social Problems) are also the most strongly coupled with IQ – suggesting a close interrelationship between these outcomes at clinical and perhaps neurobiological levels of analysis. These results highlight how combined analysis of categorical and continuous measures helps to capture the complex multifaceted psychiatric manifestations typical of neurogenetic disorders.

Second, we use 67 different dimensional variables to provide a more comprehensive and detailed picture of psychopathology than that offered by diagnostic status alone. Ranking these scales by score recapitulates prior findings that XYY syndrome is most impactful on attentional and social domains (18,44), and further refutes the already debunked but sadly tenacious association between carriage of an extra Y-chromosome and violent behavior (8). Our screen of multiple dimensions also indicated that the most severely scored symptom domains in XYY syndrome were not necessarily those that were most strongly associated with cognitive ability, adaptive functioning or caregiver strain. Most strikingly, scores on physical aggression and OCD-related domains were simultaneously amongst the least impacted in XYY and the most strongly correlated with adaptive functioning and caregiver strain. This finding highlights how the most salient features of a disorder in terms of symptom counts may not necessarily be the most functionally relevant. These cross-sectional associations provide important hints regarding the potential causal relationships between mental health, cognition, adaptive functioning and caregiver strain – informing the design of those longitudinal and interventional studies that are now needed to address causal questions.

Third, comparison of dimensional measures between XYY individuals who received their diagnoses prenatally vs. postnatally provided a detailed view of potential genetic diagnosis ascertainment bias effects in the estimation of XYY’s penetrance for psychopathology. Although most measures showed a trend towards less severe scores in prenatally vs. postnatally diagnosed individuals (especially for ASD-related features), scores in the prenatally diagnosed subgroup remained significantly elevated (median z>2) for most measured domains, and the ranking of domains remained highly stable between the subgroups. Thus, the patterning of XYY effects across different aspects of psychopathology is generally robust to potential ascertainment biases based on the timing of genetic diagnosis, and the reported capacity of XYY syndrome to impact mental health is unlikely to be solely reflective of ascertainment biases in the largely clinically recruited cohorts studied to date. However, our finding of systematic differences between pre- and postnatally diagnosed XYY individuals also argues for the importance of innovating methods for deep-phenotyping in population-based samples.

Fourth, application of WSBM to our high-dimensional behavioral data helped to distill key clinical dimensions of behavioral variability in XYY syndrome and illustrates the potentially broad utility of network analysis for parsing the characteristically complex and varied psychiatric manifestations of GDDs. Through WSBM analysis we compress >60 individual scales into 8 clusters that capture the main axes of psychopathological variation in XYY. These data-driven clusters often bring together subscales from different instruments that capture a similar aspect of symptomatology – such as the “Externalizing” cluster that includes closely related subscales from the CBCL, SDQ, Conners, SHARP and APSD. This co-clustering of related scales from different instruments demonstrates the coherence and convergent validity of WSBM clusters and suggests that it may be possible to efficiently position individual carriers within the latent behavioral space of XYY syndrome using a small handful of questionnaire items that individually capture different WSBM clusters. Moreover, interindividual variation in cluster scores shows dissociable relationships with IQ, adaptive functioning and caregiver strain. Although we lack evidence for causality, these findings suggest domains of psychopathology that could be prioritized as potential treatment targets for improving adaptive functioning (e.g. attentional impairments) and lessening caregiver strain (e.g. externalizing symptoms). Analysis of correlations between WSBM clusters reveals a “core-periphery” organization to the network of psychopathology in XYY syndrome. Inattention, externalizing behaviors, current social impairments, impulsivity and global psychopathology form the core of this network, surrounded by peripheral clusters of internalizing problems, early social impairments and dissociality which represent features that can vary more independently across XYY carriers.

Our findings should be considered in light of several study limitations. First, although our cohort represents one of the largest behaviorally characterized groups of XYY individuals to date, and captures substantial variability in presentation, it will be important to continue expanding sample sizes in future research. Second, the low prevalence of known individuals with XYY syndrome makes it impractical to recruit and deep-phenotype large samples in narrow age ranges. Reflecting this fact, our cohort age range spanned 5-25 years, which may encompass substantial age-related variation that we are unable to capture in our analysis. This limitation could be addressed when large longitudinal cohorts of individuals with XYY syndrome are assembled. Third, while we use time of XYY diagnosis as a probe for potential ascertainment bias effects, the only way to completely control for ascertainment bias is to achieve complete or fully randomized identification of XYY individuals within a population-based sampling frame. However, we note that the ranking of scales by impact remains stable between the lower and higher bias subgroups in our study – suggesting that the observed profile of relative strengths and vulnerabilities in XYY syndrome may be a stable feature. The observed profile of psychopathology in XYY syndrome from our analysis of dimensional measures is also potentially colored by the control group against which XYY scores were compared. In this initial report, we have intentionally focused on a control group of XY individuals who has been screened to verify the absence of prior psychiatric diagnoses – but an important goal for subsequent work would be use of alternative control groups (e.g. unscreened XY individuals or clinical groups with gene dosage disorders other than XYY). Fourth, the cross-sectional nature of our study prevents any causal interpretations of observed correlations between different domains of psychopathology, or between psychopathology and estimates of IQ, adaptive function, or caregiver strain. Nevertheless, we believe that characterizing such correlations is an important first step that helps to prioritize the necessarily more targeted study designs required to address causal questions. Fifth, our study design is also unable to resolve the sources of variability across individuals – which are presumably either genetic, environmental, or stochastic in nature. We hope that defining the main axes of phenotypic variability that organize psychiatric manifestations of GDDs like XYY syndrome (**Fig. 4**) will help to accelerate future studies that seek the sources and biological mediators of this variability.

## Data Availability

All data produced in the present study are available upon reasonable request to the authors

## ACKNOWLEDGMENTS

This research was funded by the NIMH Intramural Research Program ((Clinical trial reg. No. NCT00001246; clinicaltrials.gov; NIH Annual Report Number, 1ZIAMH002949-04; Protocol number: 89-M-0006).

---

**Table S1. Multidimensional Assessment of Psychopathology in XYY Syndrome**. Descriptive and analytic statistics are provided for each of 67 distinct scales derived from 12 questionnaires. Columns are as follows: Questionnaire name and abbreviation; Scale name; Unique variable name combining scale and questionnaire; the number of participants in XYY and XY groups with available measures for each variable (we confirmed that age was matched between XYY and XY groups for each variable); the mean scales variable score (mean Z) and its standard deviation (s.d.) in XYY; the coefficient (mean ΔZ) and coefficient standard error (s.e.m.) for the difference in scaled variable score in postnatally diagnosed vs. prenatally diagnosed XYY individuals; the coefficient (mean ΔZ) and coefficient standard error (s.e.m.) for the difference in scaled variable score in prenatally diagnosed XYY individuals vs. XY controls; the standardized regression slopes (β-standardized) and slope standard errors (s.e.m.) for each scale as a predictor of variation in IQ, Vineland Adaptive Behavior Composite Score (VABC) and total Caregiver Strain Questionnaire Score (CSQ).

